# Increased resting-state functional connectivity within theta and alpha frequency bands in dysphoria: Towards a novel measure of depression risk

**DOI:** 10.1101/2020.09.22.20199281

**Authors:** Carola Dell’Acqua, Shadi Ghiasi, Simone Messerotti Benvenuti, Alberto Greco, Claudio Gentili, Gaetano Valenza

**Author notes:** **Corresponding author**: Carola Dell’Acqua, Department of General Psychology, University of Padua, Via Venezia, 8 - 35131 Padua, Italy. These authors contributed equally to this work (Carola Dell’Acqua, Shadi Ghiasi).

## Abstract

**Background:** The understanding of neurophysiological correlates underlying the risk of developing depression may have a significant impact on its early and objective identification. Research has identified abnormal resting-state electroencephalography (EEG) power and functional connectivity patterns in major depression. However, the entity of dysfunctional EEG dynamics in dysphoria is yet unknown.

**Methods:** 32-channel EEG was recorded in 26 female individuals with dysphoria and in 38 age-matched, female healthy controls. EEG power spectra and alpha asymmetry in frontal and posterior channels were calculated in a 4-minute resting condition. An EEG functional connectivity analysis was conducted through phase locking values, particularly mean phase coherence.

**Results:** While individuals with dysphoria did not differ from controls in EEG spectra and asymmetry, they exhibited dysfunctional brain connectivity. Particularly, in the theta band (4-8 Hz), participants with dysphoria showed increased connectivity between right frontal and central areas and right temporal and left occipital areas. Moreover, in the alpha band (8-12 Hz), dysphoria was associated with increased connectivity between right and left prefrontal cortex and between frontal and central-occipital areas bilaterally.

**Limitations:** All participants belonged to the female gender and were relatively young. Mean phase coherence did not allow to compute the causal and directional relation between brain areas.

**Conclusions:** An increased EEG functional connectivity in the theta and alpha bands characterizes dysphoria. These patterns may be associated with the excessive self-focus and ruminative thinking that typifies depressive symptoms. EEG connectivity patterns may represent a promising measure to identify individuals with a higher risk of developing depression.

## Introduction

Depression is a globally prevalent mood disorder primarily associated with a persistent state of sadness and/or loss of interest that affects thought and behavior, showing consequent impairments in emotional and cognitive information processing (Kessler, 2012; LeMoult and Gotlib, 2018). Depressive disorders are associated with elevated rates of lifetime prevalence and chronicity and were defined as a leading cause of disease burden worldwide in 2010, with an increase of 37.5% as compared to 1990 (Ferrari et al., 2013). Accordingly, the European Pact for Mental Health and Well-Being has emphasized as key priority advancing research on the early recognition and prevention of this condition (Wahlbeck and Mäkinen, 2008). Given these premises, gaining a better understanding of neurophysiological factors underlying the risk of developing major depression could have a significant impact on its early and objective detection and treatment. A promising framework in the search of neural correlates that may predispose individuals to depression is the study of subclinical conditions. Specifically, dysphoria, or subclinical depression, is a condition characterized by elevated depressive symptoms that do not meet the criteria for a formal diagnosis of major depression – usually with respect to the frequency, duration and/or severity of symptoms (National Collaborating Centre for Mental Health, 2010; Rodrìguez et al., 2012). Dysphoria is acknowledged to considerably increase the risk of developing a full-blown major depression episode (Shankman et al. 2009; Pietrzak et al. 2013; Lee et al., 2019). Specifically, the presence of dysphoria has been reported to be the strongest risk factor for major depression onset and the leading single screening measure in a sample of female adolescents across a 4-year follow- up (Seeley et al., 2009). Besides, studying dysphoria offers the advantage to analyze neurophysiological measures associated with the risk of depression without any alterations produced by the use of antidepressant medications.

Non-invasive neuroimaging techniques, including functional magnetic resonance imaging (fMRI) and electroencephalography (EEG), have been extensively used to study resting-state alterations of brain function in depression. The resting-state paradigm reflects endogenous intrinsic brain organization and is believed to convey valuable information on how distinct brain structures communicate (Deco et al., 2011). Neural activity assessed with EEG is mostly quantified by applying time series analysis (e.g., Fast Fourier Transformation) to estimate the power spectrum of the signal within five main frequency bands (i.e., delta, 1-3 Hz; theta, 4-8 Hz; alpha, 8-12 Hz; beta, 13-25 Hz; gamma, 30-100 Hz). Activity in the theta and alpha bands was suggested to be inversely related to the level of cortical activation and, consequently, it is considered markers of the idling brain (Grin-Yatsenko et al., 2010). EEG studies point to both distributed and local connectivity disturbances (for a comprehensive review see: de Aguiar Neto and Rosa, 2019).

With regard to depression-related spectral alterations, there is a consensus agreement for higher resting-state alpha activity in parietal and occipital sites in individuals with depression (Damborska et al., 2020; Grin-Yatsenko et al., 2009; Grin-Yatsenko et al., 2010; Jaworska et al., 2012; Kemp et al.,2010; Olbrich and Arns, 2013). With respect to anterior regions, resting-state brain asymmetry in alpha power, an index of the balance between cortical activity recorded at left and right frontal scalp sites, has been widely investigated in relation to emotion and depression (e.g., Allen and Reznik, 2015; Allen and Cohen, 2010; Allen et al., 2004, Henriques and Davidson, 1991). Specifically, during resting-state, a stable pattern of increased alpha band (8-12 Hz) power (reflecting reduced cortical activity) in the left anterior scalp sites compared to the right side was observed in depression (e.g., Allen and Cohen, 2010; Coan et al., 2006; Davidson, 1998). Such resting assessments have been suggested to represent a stable biomarker of depression and are often interpreted in light of the dispositional model of motivation and affective style. The model proposes that individuals have trait-like dispositions for affective responses of approach (greater left frontal activity) or withdrawal (greater right frontal activity), irrespectively of the demands of the situation (Allen and Cohen, 2010; Coan et al., 2006; Davidson, 1998). In addition to alpha frequency band changes, depression was associated with a higher power in the theta frequency band (4-8 Hz) in parietal and occipital regions (Grin-Yatsenko et al., 2009, Grin-Yatsenko et al., 2010) and in subcortical limbic structures (Damborska et al., 2020). Moreover, most studies that investigated theta band in depression have focused on its relation to treatment outcomes (Arns et al., 2015). Specifically, greater frontal theta activity within the anterior cingulate cortex, also defined as midline theta, has been associated with a favorable treatment outcome in depression (Pizzagalli et al., 2001). Conversely, an excess of theta band widespread in the cortex has been associated with non-response to antidepressant treatments (Knott et al., 1996; Olbrich et al., 2015).

Although resting-state EEG spectral activity has been associated with depression-related affective dysregulations, inconsistent results have emerged, raising criticisms about the effective value of this methodology to identify potential biomarkers (for a recent meta-analysis see van der Vinne et al. 2017). As a matter of fact, within the last decades, it has become clear that depressive symptoms are associated with a high inter-subject variability of neurophysiological signatures. Moreover, pathophysiological changes seem to consistently occur across brain regions rather than to specific local alterations (Kaiser et al., 2015; Mulders et al., 2015). Hence, examining connections across distal brain regions provides a better understanding of the underlying mechanisms of depressive symptoms. Specifically, the estimation of the dynamical interactions among simultaneously recorded brain signals allows the identification of abnormal connectivity patterns associated with pathologies. The estimation of synchronous activity can be quantified through the identification of brain regions that have a correspondent frequency, phase or amplitude of correlated activity, namely functional connectivity. Functional connectivity indicates the temporal dependency between spatially distant neurophysiological events without, however, determining the direction of the influence that one region may have on another (Friston, 2011).

A suitable and commonly applied functional connectivity measure is EEG mean phase coherence, a correlation coefficient that estimates the consistency of relative amplitude and phase between pair of signals in each frequency band (Bowyer, 2016; Srinvasan et al., 2007). Although comprehensive evidence on resting-state functional connectivity changes in depression is still lacking, there is some support for increased alpha and theta coherence in long-distance connections (Leuchter et al., 2012; Fingelkurts et al., 2007). Medication-free individuals with depression presented higher theta and alpha coherence between frontopolar and temporal or parieto-occipital regions and higher beta coherence primarily in connections within and between electrodes overlying the dorsolateral prefrontal cortical or temporal region (Leuchter et al., 2012). Similarly, higher alpha posterior long-distance connections and lower theta left and anterior long-distance connections were observed in individuals with depression, compared to controls. Also, depression was globally associated with greater and stronger short-distance connections in the left hemisphere and long-distance connections in the right hemisphere (Fingelkurts et al., 2007). Further, heightened prefrontal connectivity within the alpha frequency between the subgenual and the left medial and dorsolateral prefrontal cortex has been reported in depression (Olbrich et al., 2014). Another study found concurrent higher connectivity in the alpha frequency band within three modules in bilateral posterior and anterior regions in participants with depression, when compared to healthy controls (Fingelkurts, 2017). These regions were matched to modules of the default mode network (DMN), a distributed resting-state network associated to self-referential thinking and rumination (Fingelkurts, 2017; Raichle, 2015). These findings are consistent with a series of fMRI studies that reported increased functional connectivity within the DMN in depression (for a review see: Kaiser et al., 2015; Mulders et al., 2015).

While most studies have examined neural correlates of major depression, to the best of our knowledge, no studies have yet provided a comprehensive EEG characterization in dysphoria. To this end, the present study aimed to investigate whether EEG spectral features and coherence patterns could embody a measure of risk of developing depression, instead of representing a mere correlate of the clinical depression state. Particularly, quantitative electroencephalography analysis, namely, power spectral estimation and functional connectivity mapping, were applied with the following hypotheses: (1) dysphoria increases spectral power in the theta and alpha frequency bands, predominantly in posterior cortical areas, (2) dysphoria lowers cortical activity in the left anterior scalp sites compared to the right side, indexed by higher alpha asymmetry, and (3) dysphoria increases functional connectivity within the theta and alpha frequency bands in distributed anterior and posterior regions as compared to the control group.

## Methods

In this section, the experimental protocol and the acquisition procedure are described. Then, details on signal acquisition, pre-processing, data reduction and statistical analysis are provided. A flowchart of the analysis of the study is illustrated in Figure 1.

**Figure 1.**
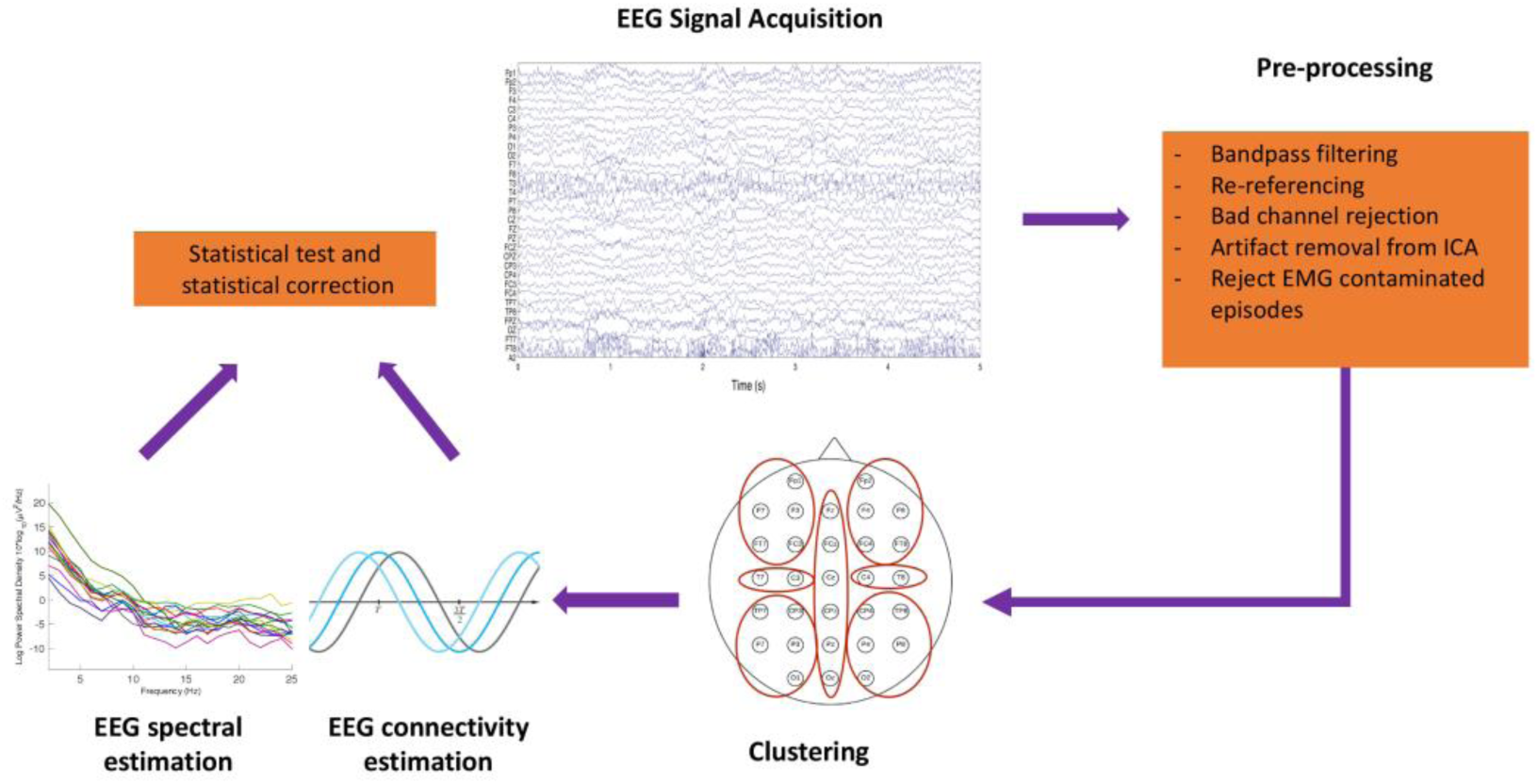
EEG data analysis pipeline.

### Participants

The present study was conducted within an extensive research project, and most of the participants’ data have also been described in previous publications (Mennella et al., 2015; Messerotti Benvenuti et al., 2017a). However, an innovative approach to data analysis has been applied in the present study.

A total of 64 female undergraduate students voluntarily took part in the study. The enrolled participants were medically healthy and free from psychotropic medication, as assessed with an ad-hoc anamnestic interview. To identify individuals with depressive symptoms, participants completed a paper-and-pencil version of the Beck Depression Inventory-II (BDI-II (Beck et al., 1996, Italian version by Ghisi et al., 2006). The BDI-II is a valid and reliable self-report questionnaire that evaluates the severity of depressive symptoms in the past two weeks and is composed of 21 items. Responses are given on a four-point (0–3) Likert scale and scores ranging from 0 to 63, with higher scores indicating more severe depressive symptoms. A score of 12 has been reported as the optimal cut-off score to discriminate individuals with and without clinically significant depressive symptoms in the Italian population (Ghisi et al., 2006). To confirm the presence of dysphoria and to exclude individuals with major depression, dysthymia or bipolar disorder, each participant scoring equal to or greater than 12 on the BDI-II was administered the mood episode module (module A) of the Structured Clinical Interview for the DSM-IV Axis I (SCID-I, First et al., 1997; Italian version by Mazzi et al., 2000). The module A of the SCID-I was administered by a trained psychologist who had previous experience with administering structured clinical interviews. Participants who scored equal to or greater than 12 on the BDI-II and had at least two current depressive symptoms, at least two weeks in duration, without meeting the diagnostic criteria for major depression, dysthymia or bipolar disorder, were assigned to the group with dysphoria (*n* = 26; age, mean (M) = 21.2, Standard Deviation (SD) = 1.8; BDI-II score, M = 16.3, SD = 4.5). The module A of the SCID-I was not administered to two of the 26 participants included in the group with dysphoria.

Participants scoring equal to or less than 8 on the BDI-II, were assigned to the group without dysphoria (*n* = 38; age, M = 22.3, SD= 2.2; BDI-II score, M= 3.2, SD=4.5). In order to ensure separation between groups with dysphoria and without dysphoria, participants who scored between 10 and 11 on the BDI-II were excluded from the present study. Participants with and without dysphoria were simultaneously recruited during the same study period. The present study was conducted with the adequate understanding and informed written consent of the participants in accordance with the Declaration of Helsinki. The study was approved by the local Ethics Committee, University of Padua (prot. No. 1407), and written informed consent was obtained from each participant enrolled in the study.

### Procedure

Prior to the experimental session, participants were required to avoid alcohol consumption the day before and to avoid caffeine and nicotine the same day of the appointment. Upon arrival at the laboratory, participants signing the informed consent were first administered a demographic interview and the BDI-II. Participants with a score equal to or greater than 12 were administered the mood episode module (module A) of the SCID-I by a trained psychologist. Then, participants were seated in a dimly lit, sound-attenuated room. After sensors were attached, EEG was recorded under eyes-open resting-state conditions over a 4-minute period for each participant.

#### Electroencephalographic recordings

The EEG was recorded from 32 scalp positions using an Electro-Cap (Electrocap, Inc.) with tin electrodes. The EEG sites were Fp1, Fpz, Fp2, F7, F3, Fz, F4, F8, FT7, FC3, FCz, FC4, FT8, T3, C3, CZ, C4, T4, TP7, CP3, CPz, CP4, TP8, P7, P3, Pz, P4, P8, O1, Oz, O2 and A2 (right mastoid), all referenced online to A1 (left mastoid). To control for eye-movements and eye-blinks, both vertical and horizontal electro-oculograms (EOGs) were recorded, using a bipolar montage. The electrodes pairs were placed at the supra- and suborbit of the right eye and at the external canthi of the eyes, respectively. All electrode impedances were kept below 5 kΩ. The signal was amplified with Neuroscan Synamps (El Paso, TX, USA), bandpass filtered online at 0.1-70 Hz, digitized at 500 Hz (16-bit AD converter, accuracy 0.034 μV/bit), and stored on to a Pentium II computer. ECG recording was also conducted to investigate heart rate variability (data published in Messerotti Benvenuti et al., 2015a; Messerotti Benvenuti et al., 2015b).

#### Data preprocessing

The EEG signal was acquired with a sampling frequency of 500 Hz and re-referenced to an average reference. The EEG was filtered offline using a band-pass Butterworth filter between 0.5 Hz and 45 Hz and manually corrected by removing the components associated with artifacts (e.g., blink artifacts) using the independent component analysis (ICA) as implemented in EEGLAB (Delorme and Makeig, 2004; Jung et al., 2000). Finally, all the signals were inspected visually to remove all the remaining artifacts related to movement or other sources.

In order to perform statistical analysis on EEG data, a cluster-based approach has been applied to control over type I error rate arising from multiple comparisons across electrodes and time points (Henriques and Davidson, 1991). Seven clusters were obtained by the grouping of 30 channels (the mastoids were excluded from the analysis), according to our regions of interest.

- Cluster 1 (C1): F7, FP1, F3, FT7, FC3
- Cluster 2 (C2): F8, FP2, F4, FT8, FC4
- Cluster 3 (C3): T3, C3
- Cluster 4 (C4): T4, C4
- Cluster 5 (C5): P7, P3, O1, CP3, TP7
- Cluster 6 (C6): P8, P4, O2, TP8, CP4
- Cluster 7 (C7): FZ, CZ, PZ, FCZ, CPZ, OZ

In each cluster a new time series is achieved as a result of the average among the channels.

## Data analysis

### Quantification of neural activity and neural network

Spectral analysis for quantifying the neural activity was applied. Particularly, the analysis involved the computation of time-frequency representation of each cluster time series using the Welch periodogram, with a Blackman window of 4 s and 75% of overlap. This spectrum was then integrated within two bands of interest (alpha and theta; Babiloni et al., 2016). Consequently, power spectral estimates in the alpha and theta frequency bands were calculated for each subject at each of the seven clusters of interest. Asymmetry score was calculated for total alpha power by subtracting the natural log-transformed scores for each homologous left and right pair (i.e., ln(Right) – ln(Left)) in anterior (FP1& FP2, F7 & F8, F3 & F4) and in posterior (P3 & P4, T5 & T6, 01 & 02) regions. Then, average asymmetry scores were calculated for each region as the average of the asymmetry scores obtained from each pair of electrodes (see also Schleiger et al., 2014)). Higher asymmetry scores reflect greater left relative to right cortical activity (Coan et al., 2006).

For the quantification of brain network, a functional connectivity analysis was applied to measure the neuronal synchrony between two regions. One way to manifest the neuronal signal synchrony is to calculate the phase synchronization, which has the advantage of being time-resolved and only sensitive to the phases irrespective of the amplitudes of two signals. The concept of phase synchronization in physics is defined as locking of the phases of two oscillators. In case of complete synchronization, the two oscillators are coupled sufficiently strong. Mean phase coherence (MPC) has been introduced as a suitable measure of phase synchronization in biological time series due to its robustness against noise (Mormann et al., 2000; Quiroga et al., 2002). Spatial and temporal shifts in synchronization that appear to be strongly related to pathological activity may be reflected in the MPC (Mormann et al., 2000). In order to compute the phase synchronization between two time series *(x(t), y(t))*, their instantaneous phases *φ(x(t))* and *φ(y(t))* are determined after calculating the analytical signal from the Hilbert transform (Kreuz, 2011). By subtracting the instantaneous phases *(Δ(φ))*, the relative phase difference is obtained and the MPC is calculated as follows:

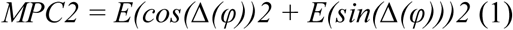

where E is the expectation operator. The value of MPC is close to 1 where there is a high phase synchronization between two time series, in our case the two EEG channels, and is close to zero where there is a weak synchronization (Valenza et al., 2018). MPC was estimated for each pair of channels among the 7 selected regions for each subject.

### Statistical analysis

In order to analyze potential between-group differences, a statistical comparison using the nonparametric Wilcoxon Mann-Whitney test was conducted. Multiple tests were performed to compare each cluster power spectrum and each connectivity in both theta and alpha bands. The obtained p-values were then adjusted by applying the false discovery rate (FDR) for multiple hypothesis testing (Benjamini et al., 2006).

Moreover, in order to investigate the relationship between the BDI-II scores and the brain activity, a correlation analysis based on Spearman correlation coefficient was estimated between each subject’s BDI-II score and the corresponding EEG power computed in the band of interest (i.e., the alpha and theta frequency bands).

## Results

### Spectral estimates

Both groups showed a similar topographic distribution of the power spectrum for both frequency bands (Fig. 2). Theta power topographic distribution showed a high value in the posterior region that decreased in the central-temporal area and slightly increased in the frontal regions. The alpha power topographic distribution showed a high activity in the midline and posterior regions, which significantly decreased in temporal area.

**Figure 2.**
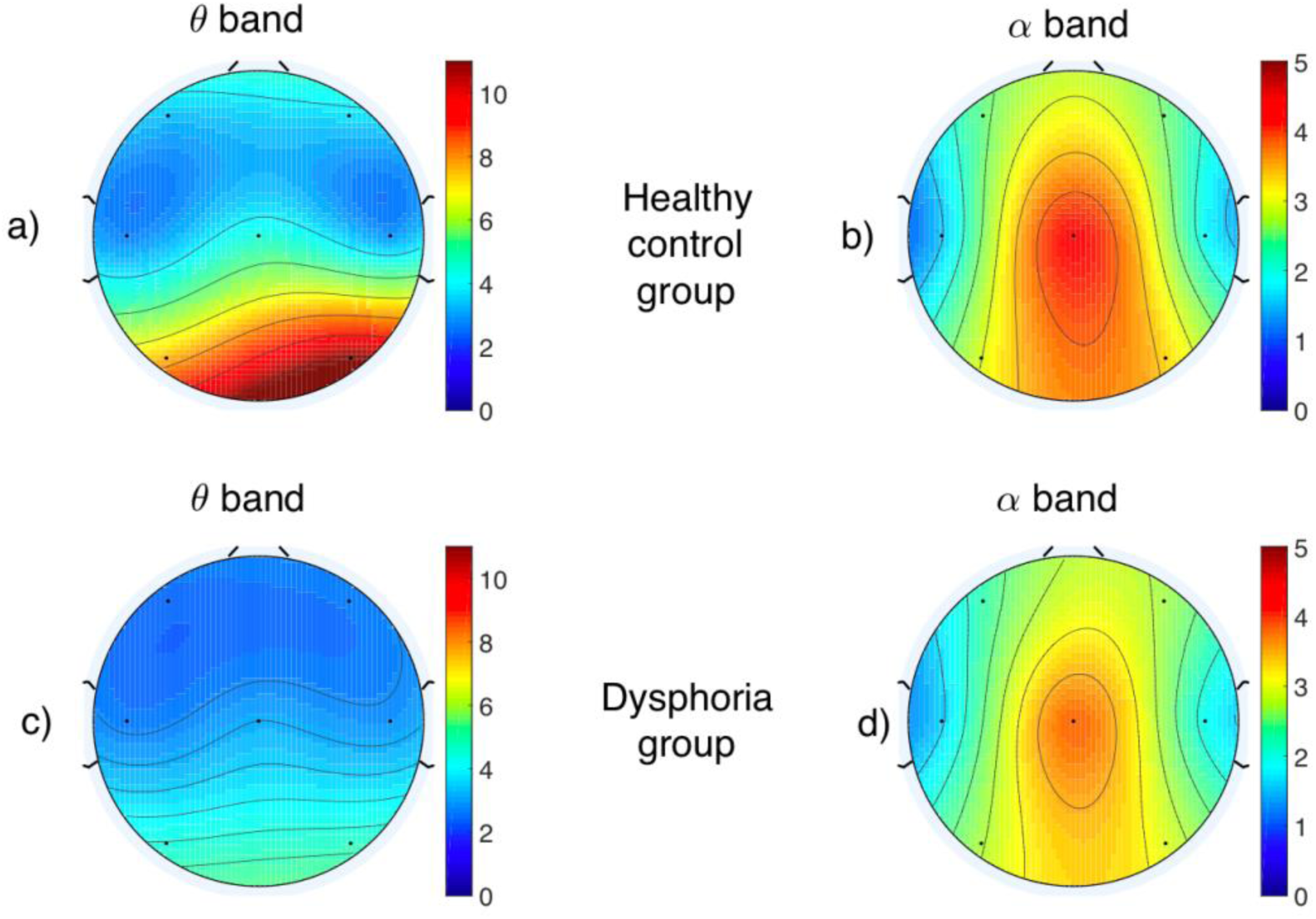
Topographic map of the EEG power spectrum computed in both the theta and alpha frequency bands.

The similar distribution between the two groups were confirmed by the statistical comparison. There was no significant difference in spectral measures between the group with dysphoria and the control group in any of the scalp regions (Fig. 3). Likewise, the analysis revealed no significant differences among the two groups in the alpha asymmetry index of both frontal and posterior regions (all *Ps* > 0.05).

**Figure 3.**
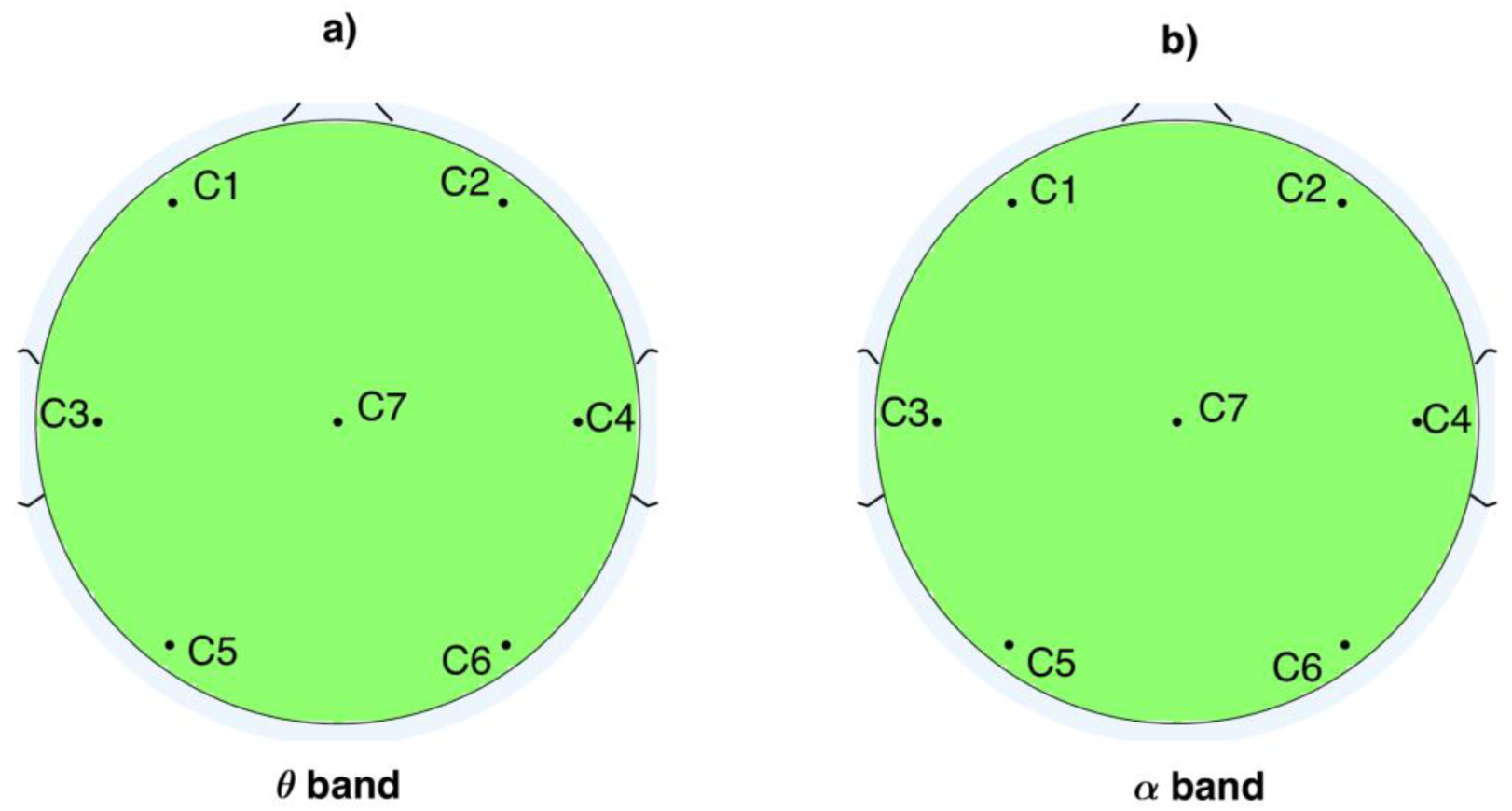
(Panel a) P-value topographic maps obtained from Wilcoxon tests on EEG power spectral density in the theta band. (Panel b) P-value topographic maps obtained from Wilcoxon tests on EEG power spectral density in the alpha band. The color map goes from blue (corresponding to the p-value of 0.02), when there is a significant increase of the power in individuals with dysphoria compared to healthy control, to red (corresponding to the p-value of 0.02) when a significant decrease of power in individuals with dysphoria has been observed. Green indicates no difference between the two groups.

### Functional Connectivity analysis

Statistically significant changes of cortical connection in individuals with dysphoria among the seven brain regions for alpha and theta bands were observed (Fig. 4). More specifically, a strong increase in the neural connectivity of the group with dysphoria compared to the control group in theta band was found between two pairs of regions, right frontal with fronto-central (C2 and C7, *p* = 0.047) and right temporal with left occipital (C4 and C5, *p* = 0.042) (Fig. 4, panel a). Concerning the alpha band, we observed a significant increase of MPC index in individuals with dysphoria among five pairs of regions. Particularly, an increase in synchronization was found between the right and left prefrontal cortices (C1 and C2, *p* = 0.010) and between frontal and central-occipital areas bilaterally (C5 and C1, *p* = 0.040; C6 and C2, *p* = 0.009; C7 and C2, *p* = 0.027; C7 and C6, *p* = 0.028). Additionally, dysphoria was associated with a significant decrease in functional connectivity between the right frontal and temporal areas in the alpha band (C2 and C4, *p* = 0.021) (Fig. 4, panel b).

**Figure 4.**
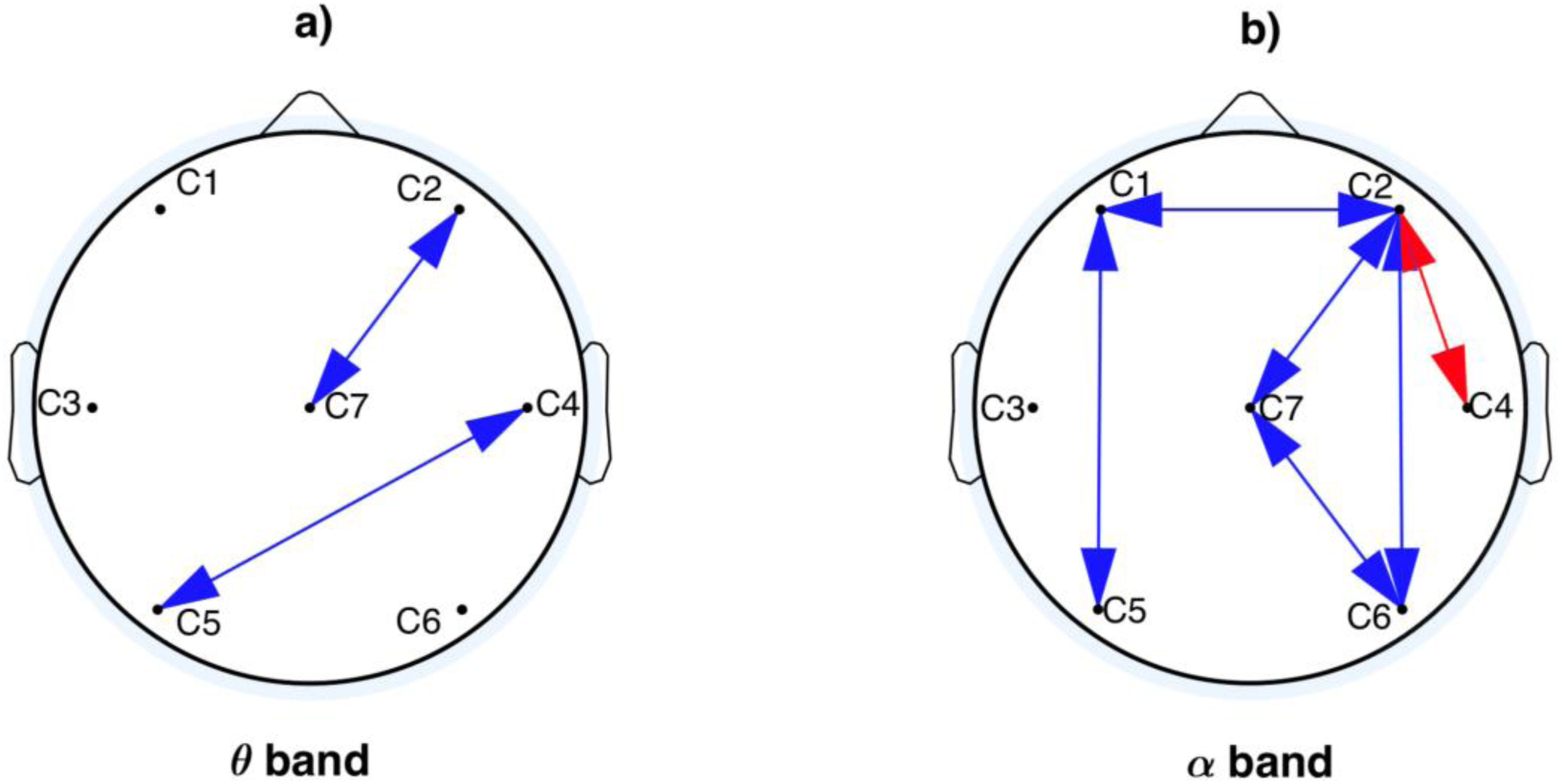
(Panel a) Mean phase coherence significant decrease (red arrows) and significant increase (blue arrows) in individuals with dysphoria compared to healthy controls among 7 regions of interest in the theta band. (panel b) Mean phase coherence significant decrease (red arrows) and significant increase (blue arrows) in individuals with dysphoria compared to healthy controls among 7 regions of interest in the alpha band. The p-values are corrected for multiple comparison.

### Spearman Correlation analysis

Figure 5 depicts the results of a group-wise Spearman’s correlation analysis between the EEG power spectrum estimated in the theta and alpha bands and the BDI-II scores in terms of correlation coefficient and p-value.

**Figure 5.**
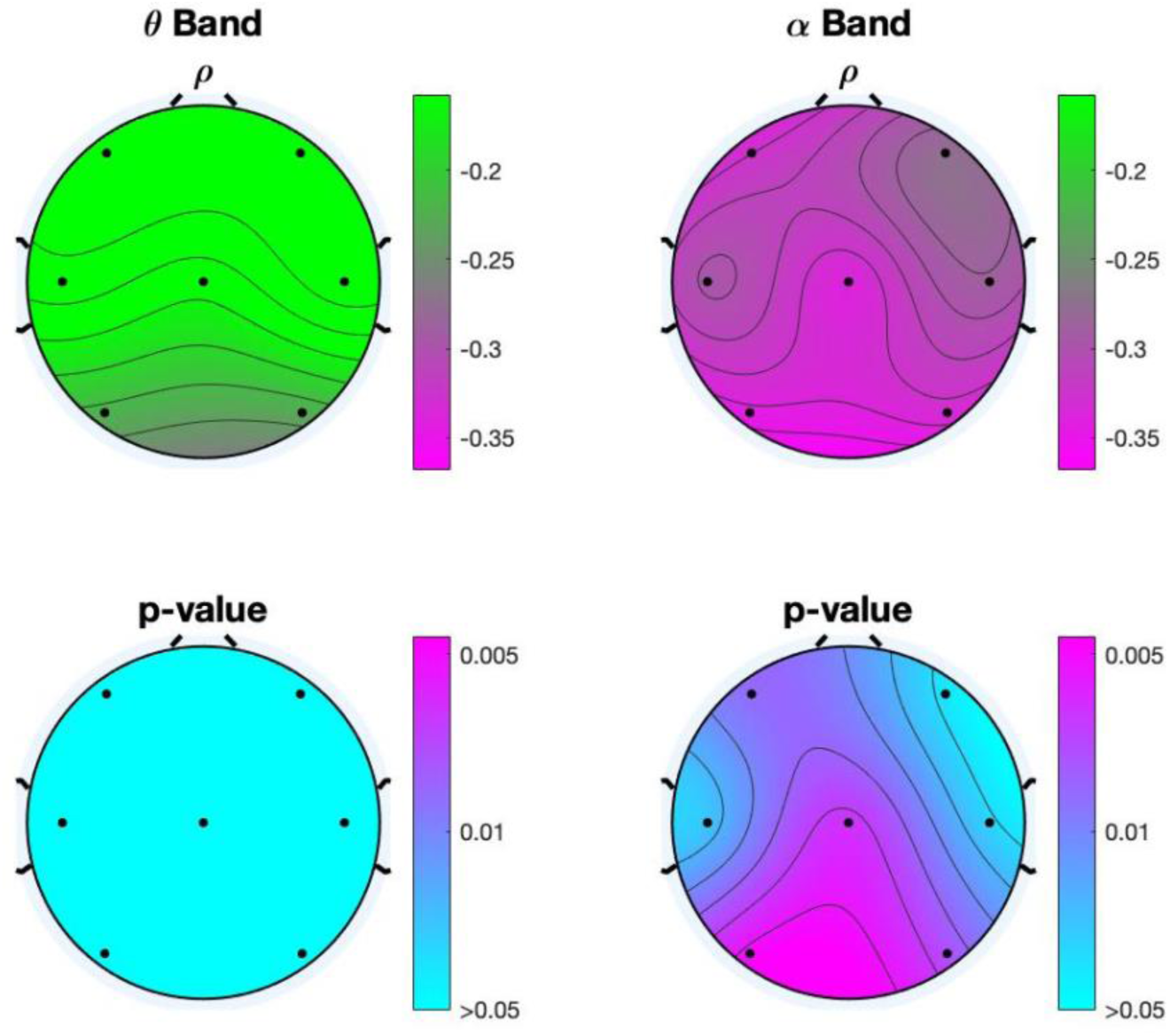
Spearman correlation coefficient (top) and related p-values (bottom) calculated between the BDI-II scores of all subjects and the related EEG power in the theta and alpha frequency bands.

Concerning the theta band, no significant correlation (*all Ps* > 0.05) was found between the considered psychometric measure and the EEG activity. On the other hand, a significant negative correlation between EEG alpha power spectrum and BDI-II scores was observed in several brain regions. Specifically, a negative Spearman’s correlation (index in the ranges of - 0.327, -0.344) was associated with the posterior (C5, C6) and midline (C7) brain areas and the frontal left region (C1).

## Discussion

Resting-state EEG was analyzed to determine whether dysphoria may be effectively characterized by spectral and functional connectivity changes with respect to healthy neural patterns. The aim was to identify effective biomarkers of subclinical conditions, whose underpinning functional neural patterns are yet unclear (Fischer et al., 2016). This may also contribute to its early and objective identification, to an effective treatment monitoring, therefore reducing the individual’s risk of developing major depression. Based on previous studies, with respect to healthy state, it was hypothesized that dysphoria may be characterized by (1) an increase of spectral power in the theta and alpha frequency bands, predominantly in posterior cortical areas, (2) lower cortical activity in the left anterior scalp sites compared to the right side, indexed by higher alpha asymmetry and (3) increased functional connectivity within the theta and alpha frequency bands in distributed anterior and posterior regions compared to the control group.

With respect to power spectral estimates, conversely to what hypothesized, no significant differences were detected between the group with dysphoria and the control group in alpha and theta powers. Furthermore, the correlation analysis revealed a significant inverse correlation between alpha power and BDI-II scores, predominantly in midline and posterior regions bilaterally. In other words, the alpha power distribution at a group level differed as a function of depression scores, with a higher alpha power associated with lower BDI-II scores. This finding is in line with previous studies that reported similar findings (Volf and Paaynkova, 2002; Zoon et al., 2013; Jian et al., 2016), observing an inverse correlation between alpha power and degree of depressive symptoms, although opposite findings have also been reported (Damborska et al., 2020; Grin-Yatsenko et al., 2009; Grin-Yatsenko et al., 2010). Considering that alpha power is an inverse index of cortical activity and consequently as a marker of the idling brain (Grin-Yatsenko et al., 2010), decreased alpha power in the posterior brain regions may reflect higher neuronal excitability. Thus, it could be hypothesized that reduced alpha power in relation to higher BDI-II scores may reflect the sustained state of stress associated with the presence of depressive symptoms (Heller et al., 1998).

The finding that the two groups did not differ in the measure of frontal alpha asymmetry is not in line with a series of studies that associated this index with depression (e.g., Allen and Cohen, 2010; Coan et al., 2006; Davidson, 1998). On the other hand, the effective value of resting frontal alpha asymmetry as a potential biomarker for depression was challenged by the emergence of inconsistent results (for a recent meta-analysis see van der Vinne et al., 2017). Indeed, recent findings suggest that task-related activity, instead of resting-state, is a more sensitive measure of depression (Coan et al., 2006). Moreover, affective dispositions dysregulations, associated with frontal alpha asymmetry, have been suggested to be more pronounced in response to emotional stimuli, since the emotional demands of the context highlight the affective deficit (Coan et al., 2006; Mennella et al., 2015; Messerotti Benvenuti et al., 2019; Stewart et al., 2014).

Brain coherence may effectively be used to characterize resting-state activity, as it reflects the functional organization of brain networks that subserve a range of processes associated with depression, such as self-referential processing and emotional saliency (Deco et al., 2011). In line with what hypothesized, strengthened functional connectivity in distributed neural networks was observed in participants with dysphoria compared to healthy controls. Specifically, in the theta band these differences were significant between two pairs of regions, right frontal and central areas (C2 and C7) and right temporal and left occipital areas (C4 and C5) (Fig. 3, panel a). Concerning the alpha band, an increase in synchronization was found among the right and left prefrontal cortex (C1 and C2) and between frontal and central-occipital areas bilaterally (Fig. 3, panel b). Moreover, results showed a strong decrease in neural coherence of individuals with dysphoria in the alpha band between the right frontal and temporal areas (C2 and C4). These findings are in line with previous studies showing heightened intrinsic functional connectivity in medication-free individuals with depression within the alpha and theta bands mainly in long-distance connections in the right hemisphere (Fingelkurts et al., 2007; Leuchter et al., 2012; Olbrich et al., 2014). The function of lower frequency oscillations (i.e., alpha and theta) is recognized to link distant neuronal assemblies together into functional working units through top-down control and, in turn, to modulate the activity of local functional units that are linked together by faster oscillations (Buzsaki and Draguhn, 2004; Leuchter et al., 2012; Klimesch et al., 2007). Moreover, the alpha band is the predominant EEG frequency of the human brain during mind-wandering and it has been positively correlated with the activation of the DMN (Laufs et al., 2003; Mantini et al., 2007; Jann et al., 2009; Fingelkurts 2017). The DMN is a network implicated in the processing of the individual’s internal emotional state and, specifically, to rumination (Raichle, 2015; Berman et al., 2014). In line with this, increased low frequencies coherence (alpha and theta bands) was associated with internally generated mental activity (i.e., rumination) in healthy participants (Andersen et al., 2009). These findings suggest that increased functional connectivity within the alpha frequency band may be associated with an excessive self-focus and rumination, which are typical depressive symptoms. Moreover, the present study not only found an increased connectivity between distant brain regions, but also within frontal regions (C1, C2). This finding is in line with a previous EEG study that reported increased connectivity within the anterior portion of the DMN, which seems to have a role in the integration of many aspects of emotional and social experience into self-relevant context information (Fingerlkurts et al., 2017). Thus, enhanced connectivity within this module may lead to increased self-focus, which is considered a key feature of depression (Northoff et al., 2007). Of note, these findings are in line with several fMRI studies that reported increased resting-state functional connectivity within the DMN in major depression and dysphoria (for a review see Kaiser et al., 2015 and Mulders et al., 2015; Philippi et al., 2015).

Although the neurophysiological mechanisms underpinning the dysregulation of brain oscillations in depression are still unknown, an interesting perspective may be given by the so-called monoamine hypothesis. The monoamine hypothesis states that depression is associated with reduced levels of the neurotransmitter serotonin in distributed synaptic transmissions (Moreno et al., 2000). Serotoninergic axons reach a wide range of brain structures and are involved, among many functions, in the modulation of brain functional connectivity. Serotonin modulates functional connectivity through its effects on inhibitory cells that regulate rhythmic oscillations in the brain. Specifically, serotoninergic projections from the medial septal area inhibit hippocampal theta oscillatory synchrony and projections from the raphe nuclei to the thalamus inhibit alpha synchrony (Mu and Has, 2010). Thus, while depression is characterized by reduced serotoninergic projections (Moreno et al., 2000), this neurochemical modulation may have a direct impact on the increased synchronization within networks that are involved in mood regulation and affective processing, known to be directly regulated by subcortical limbic structures (Kudina et al., 2004).

The current study presents strengths that distinguish it from other work in this area. First, individuals with medical diagnoses (e.g., neurological, cardiovascular diseases) were excluded, as well as those under medications, thus avoiding potential confounding effects on intrinsic brain functional connectivity. Second, to the best of our knowledge, this is the first study that measured EEG coherence sin dysphoria, providing an insight into the potential mechanisms that may predispose to full-blown depression onset.

### Limitations

In interpreting our results, some limitations should be also acknowledged. First, all participants belonged to the female gender, as well as their relatively young age may not allow generalization of the findings. However, it has to be noted that dysphoria is characterized by a high female preponderance (Meeks et al., 2011; Rodrìguez et al., 2012). Also, the young age of the participants corresponds to the life phase between late adolescence and early adulthood and was selected because it is a sensitive period for the development of depressive symptoms (Grano et al., 2015). Second, functional connectivity was examined with the MPC measure, which indicates the functional association between EEG time-series data within specific frequency bands. While MPC does not allow the causal and directional assessment between brain regions, it provides reliable connectivity estimates without the need for so-called surrogate data analyses, being also robust to dynamical noise (Mormann et al., 2000; Quiroga et al., 2002).

## Conclusions

Dysphoria is characterized by a higher functional brain connectivity in the alpha and theta bands, compared to healthy controls. These patterns may be associated with typical depressive symptoms like excessive self-focus and ruminative thinking. Dysfunctional EEG functional connectivity patterns may embody quantitative measures allowing for an early identification of depressive risk, and an effective and objective treatment monitoring. Future studies are warranted to explore whether other populations that have a higher risk to develop depression (i.e., remitted patients and individuals with familiarity) are characterized by EEG dysfunctional connectivity.

## Data Availability

All data and MATLAB code will be made available to researchers upon reasonable request.

